# The impact of climate and demographic changes on future chikungunya burden and the potential role of vaccines: a mathematical modelling study

**DOI:** 10.64898/2026.02.16.26346397

**Authors:** Oscar Cortes-Azuero, Emilie Finch, Gabriel Ribeiro dos Santos, Ellie Sutcliffe, Danny Scarponi, Sadie Ryan, Henrik Salje

## Abstract

**Background:** Chikungunya virus (CHIKV) is an *Aedes* transmitted arbovirus. Demographic changes coupled with the expanding footprint of the mosquito from climate change have the potential to shift the global burden from the virus.

**Methods:** Here we use projections of human demography and *Aedes* mosquitoes’ distribution to estimate baseline and future burden from CHIKV under different climate change scenarios in 178 countries. We then estimate the potential of vaccines to mitigate the growing burden.

**Findings:** We found that under RCP2.6 (an optimistic climate change scenario), the global population at risk from CHIKV will increase by 30.2% to 5.4 billion individuals. We estimated a 35% increase in annual infections, 49% increase in cases and a 128% increase in deaths. A similar impact was found under the more pessimistic RCP8.5 climate change scenario. In Europe and the Americas, the growing presence of *Aedes* will drive the growing case burden, with increases in human population size being key elsewhere. Ageing populations will result in major increases in the number of CHIKV-related deaths in all continents outside Africa. Vaccinating 50% of individuals aged 12y+ with a vaccine providing 70% protection against disease and 40% protection against infection would avert 29% of cases and 31% of deaths.

**Interpretation:** These findings highlight how climate change will expand the footprint of CHIKV circulation, while demographic changes will lead to substantially increased case burden in affected countries. Vaccines will be critical to minimising this changing global burden.

**Funding:** CEPI

## Introduction

Chikungunya virus (CHIKV) is an arbovirus spread by *Aedes* mosquitoes^1^. Infection by CHIKV can lead to severe disease and death, including long-lasting arthralgia^1^. It has been estimated that currently there are 35 million infections and 3,700 deaths per year from CHIKV^2^. The location and seasonality of *Aedes* mosquitoes is sensitive to temperature and rainfall as well as other climatic- and land use-related factors^3–5^. The changing climate is shifting the footprint covered by mosquitoes as well as the number of months in a year that the mosquito is found^6–9^. Changing temperatures can either increase or decrease the survival of *Aedes* mosquitoes, depending on the magnitude of the temperatures reached^10^. The impact from the changes in the distribution of the vector on CHIKV infection and disease risk remain unclear, although there are increasing reports of CHIKV outbreaks in locations where it has historically not been found, including in Europe and North America^11^. Further, the demography of countries is changing. In particular, there is a progressive ageing of populations, driven by longer life expectancies and reducing birth rates. Future demography, and distribution of populations within a country, will also depend on climate ^12^. Demographic changes will alter the number and age profile of the population affected by CHIKV, especially as the risk of severe disease and death is increased in older individuals, however, the implications on case burden at national and global levels remains unclear^13^.

Understanding where CHIKV is likely to circulate under different climate change projection scenarios is needed to help guide the appropriate use of medical countermeasures. In particular, vaccines are starting to become available, with two vaccines, IXCHIQ (Valneva) and Vimkunya (Bavarian Nordic), licensed by a number of regulatory bodies^14,15^. Knowing where to target the vaccines and quantifying their potential impact are needed to develop vaccine investment cases.

In order to estimate the future distribution of CHIKV infection around the world we leverage spatial modelling efforts that have used temperature and other climate predictions under different climate change scenarios to map the distribution of the two principal vectors of CHIKV, *Aedes albopictus* and *Aedes aegypti*^*6,7*^. Particularly useful are modelling efforts that have quantified both the potential spatial distribution of the vectors and the number of months that the vectors will be found in a year - thereby incorporating both spatial and seasonal changes in the range of the vector^6^. We can combine these estimates with projections of how global populations will be spatially distributed under different climate change scenarios, as well as the size and age structure of the populations (shared socioeconomic pathways, SSPs) ^16^.

We applied mathematical models to projected future distributions of *Aedes albopictus* and *Aedes aegypti* mosquitoes under different Representative Concentration Pathways (RCP) and linked these with corresponding Shared Socioeconomic Pathway (SSP) population projections. The first combined scenario pairs the RCP2.6 with the SSP2 population projection. RCP2.6 is an optimistic scenario that assumes that substantial international efforts result in significant reductions in emissions that limit global warming to below 2°C and SSP2 represents a “Middle of the Road” future with moderate fertility and mortality declines. The second combined scenario pairs RCP8.5 with SSP5. This scenario implies a high emission future with continued high reliance on fossil fuels and large global temperature increases combined with demography driven by fossil fuels, high material consumption, strong globalization and high mobility. For each scenario, we estimate the size of the human population at risk of infection in each country as well as the number of annual incident cases and deaths. We then estimate the cases and deaths averted through vaccination strategies.

## Methods

### Mosquito and human distributions

We used published estimates of the number of months that *Aedes aegypti* and *Aedes albopictus* mosquitoes are present in a year currently and in 2050 under RCP2.6 and RCP8.5 scenarios^6^. In a sensitivity analysis, we also use an alternative source for the baseline distribution of *Aedes*^*17*^. For humans, we use modelled estimates of the population distribution under SSP2 (which we link with RCP2.6) and SSP5 (which we link with RCP8.5)^16^. Taking each country with a population size of over 200,000 individuals in turn, we divided the country into 100 × 100m^2^ grid cells. We then calculated a population weighted average of the months at risk under each scenario (today, RCP2.6/SSP2 in 2050 and RCP8.5/SSP5 in 2050).

### Estimating population at risk

Using the results of a prior literature review^2^, we fit a nonparametric regression model with a smoothing parameter to whether a country has ever experienced chikungunya transmission and the mean mosquito burden as the sole explanatory variable ^18^. We then used this fitted model to estimate the effective size of the future population at risk in each grid cell in 2050 using the estimated months of mosquito presence in 2050 under the two climate change scenarios.

### Quantifying the number of infections, cases and deaths

The average annual force of infection for CHIKV has been estimated at 1.6% per year in epidemic settings and 2.1% in endemic settings^2^. We used these estimates to calculate the probability that an individual of age *a* living in country *j* under climate change scenario *k* gets infected using a catalytic model:

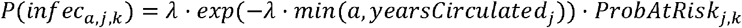

Where *λ* is the force of infection and *yearsCirculated* is the assumed number of years that CHIKV has circulated in the location (assumed to be 30 years) and *ProbAtRis k*_*j,k*_ is the effective proportion of the country at risk of CHIKV. We assume 30 years of circulation to account for transmission between 2020 and 2050. The number of infections within a country can be then calculated as:

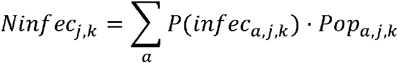

Where *Pop* _*a,j,k*_ is the size of the population of age *a* in country *j* under scenario *k* and was obtained from by International Institute for Applied Systems Analysis (IIASA) for SSP2 and SSP5 scenarios ^19^. To capture the number of cases, chronic infections and deaths following infection, we used existing estimates of the probability of disease following infection^13^.

### Isolating the independent effect

To identify the independent effects of the changing distribution of the vector, population size and age structure on CHIKV burden, we re-estimated the number of infections, cases and deaths using different sets of the data. Firstly, we used 2025 population estimates but assumed that the spatial distribution of the population at risk was given by RCP2.6 or RCP8.5. This provides a hypothetical independent effect of changing mosquito presence without demographic changes. We next used the mosquito distribution from today but used the total population size in each country from SSP2 (and separately under SSP5) but keeping the age structure the same as 2025 to identify the independent effect of population size. Finally, we used the mosquito distribution and population size from today but used the age structure from SSP2 (and separately under SSP5) to identify the independent effect of age structure.

### Quantifying the impact of vaccines

To model the impact of vaccines we considered that the force of infection will be reduced due to the partial infection blocking nature of the vaccine.

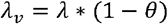

Where *λ*_*v*_ is the force of infection in a vaccinated population, *λ* is the force of infection in an unvaccinated population and *θ* is the effective reduction in the force of infection from the indirect protection generated by the vaccine. From prior work, we have found that as vaccines are not given to children (who are more likely to be susceptible), under a base case of vaccinating 50% of adults would only result in modest levels of indirect protection, we therefore consider a value of 0.05 for *θ*. We then calculated the probability that a vaccinated individual of age *a* in country *j* will be infected as:

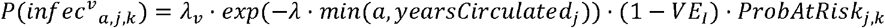

Where *V E*_*I*_ is the direct protection from infection provided by the vaccine. This assumes that the build up of immunity from natural infection prior to the introduction of the vaccine is dictated by *λ*. The unvaccinated individuals in the same population have the following probability of infection:

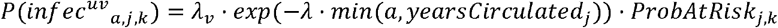

The overall number of infections in the population then becomes:

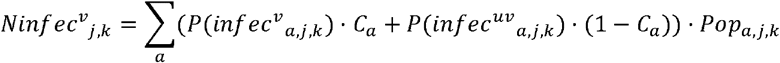

Where *C*_*a*_ is the vaccination coverage for individuals of age *a*. For epidemic settings, we assume that 20% of infections from that outbreak have already occurred by the time the desired level of vaccination coverage has been achieved.

The number of cases, chronic cases and deaths can then be derived from the number of infections, where we assumed an overall protection from disease of 70%. We conducted sensitivity analyses where we considered that (a) the vaccine was 98% effective at preventing disease; (b) the vaccine was deployed in responsive campaigns when only 1% of all cases from the outbreak have occurred in the counterfactual scenario of no vaccine use (compared to 20% of cases in the basecase); (c) coverage was 90% and (d) the vaccine was integrated into childhood immunisation so that 90% of individuals over the age of 1 years were vaccinated across endemic and epidemic at risk regions.

More details of the analytical methods can be found in the Supplementary Materials.

## Results

### Impact of climate change on CHIKV burden

We estimated the burden from CHIKV in 178 countries today and under different climate change scenarios. We found that estimates of the average predicted number of months that *Aedes aegypti* and *Aedes albopictus* are found within a location from a previous modelling study is highly correlated with the national risk of having observed CHIKV transmission (Figure 1A-B). This supports our ability to use these estimates to capture both the spatial and the seasonal impacts of climate change.

**Figure 1.**
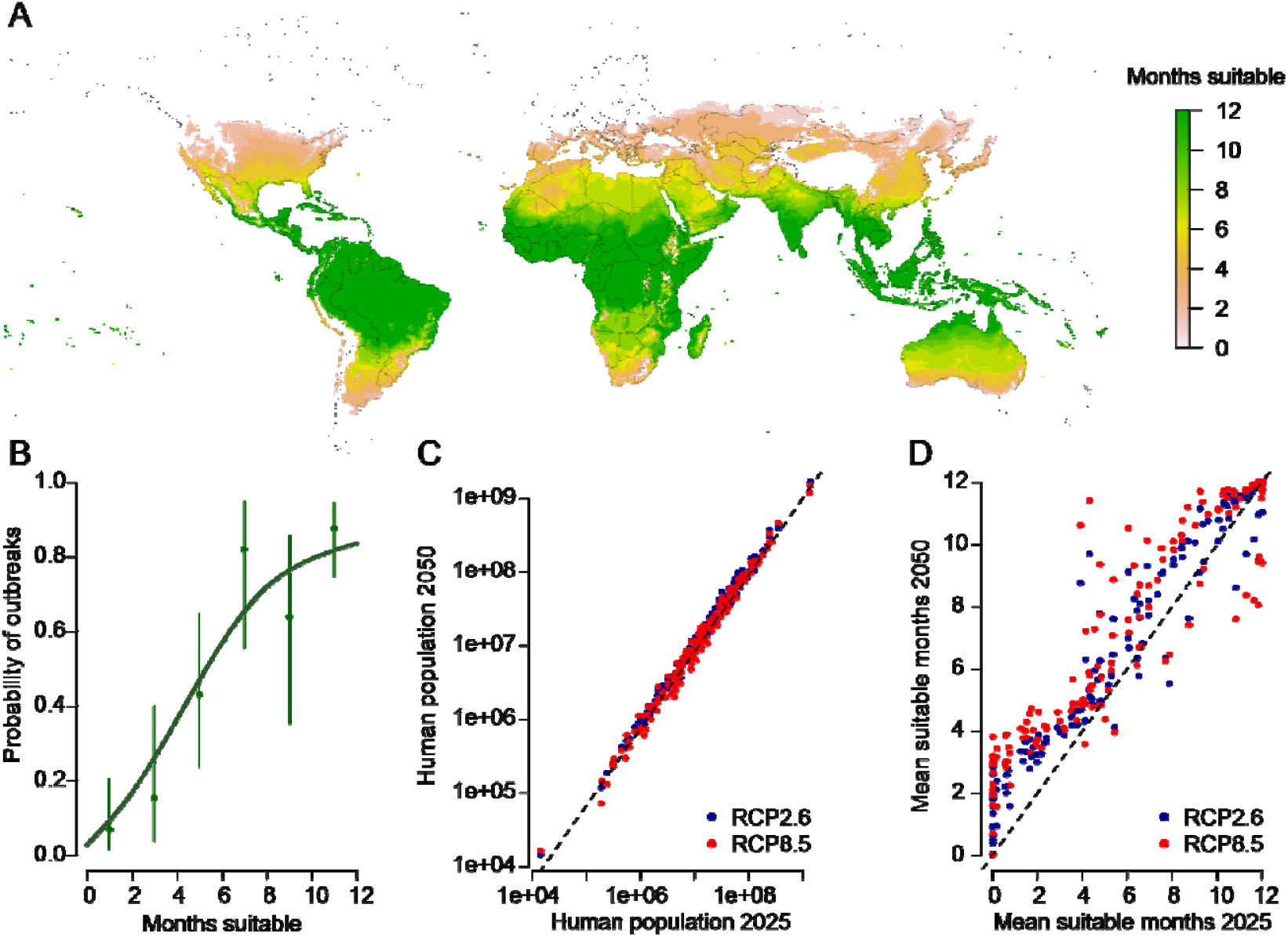
Mosquito distribution today. **(A)** Estimated number of months of *Aedes* presence today (maximum of *Aedes albopictus* and *Aedes aegypti* in each location). Derived from Ryan et al.,^6^. **(B)** Relationship between average months of mosquito presence within a country (maximum of *Aedes albopictus* and *Aedes aegypti*) and the probability that the country has ever reported CHIK transmission. The green line represents the fit of a non-parametric regression model. **(C)** Comparison between human population in 2050 and 2025 for two climate change/SSP scenarios. Each dot is the average from a single country. **(D)** Estimated mean number of suitable months for transmission in 2050 and 2025 for two climate change/SSP scenarios. Each dot is the average from a single country.

The predicted size of the populations in each country was similar across the two climate change scenarios, although the overall size of the global population in the countries analyzed was 0.6 billion greater under SSP2 in 2050 than SSP5 (8.9 vs 8.3 billion) (Figure 1C). We found heterogeneity in the predicted months of suitability for transmission in each country (Figure 1D). Under RCP2.6 we estimated that globally there will be 0.9 more months during the year that are suitable for CHIKV transmission (rising from 6.8 months to 7.7 months per year) with a similar rise expected under RCP8.5. We estimated 88% of countries are expected to see an increase in the mean months of CHIKV transmission compared to 83% under RCP8.5.

Under RCP2.6, we find that the size of the global population at risk of CHIKV infection will increase by 30.2% to 5.4 billion individuals, or 61.0% of the global population, up from 54.1% in 2025 (Figure 2A). All continents were predicted to experience a large increase in the population at risk ranging from a 16% increase in Asia to a 123% increase in Europe (Figure 2B). Under RCP8.5, the proportion of the population at risk will rise by 23.0% to 5.1 billion individuals, or 61.8% of the global population, ranging from a 7% increase in Asia to a 218% increase in Europe (Figure 2C). The few countries in both scenarios expected to see a net reduction in the proportion of the population at risk are located in Central Africa and the Middle East where temperature rises will exceed those optimal for *Aedes* survival.

**Figure 2.**
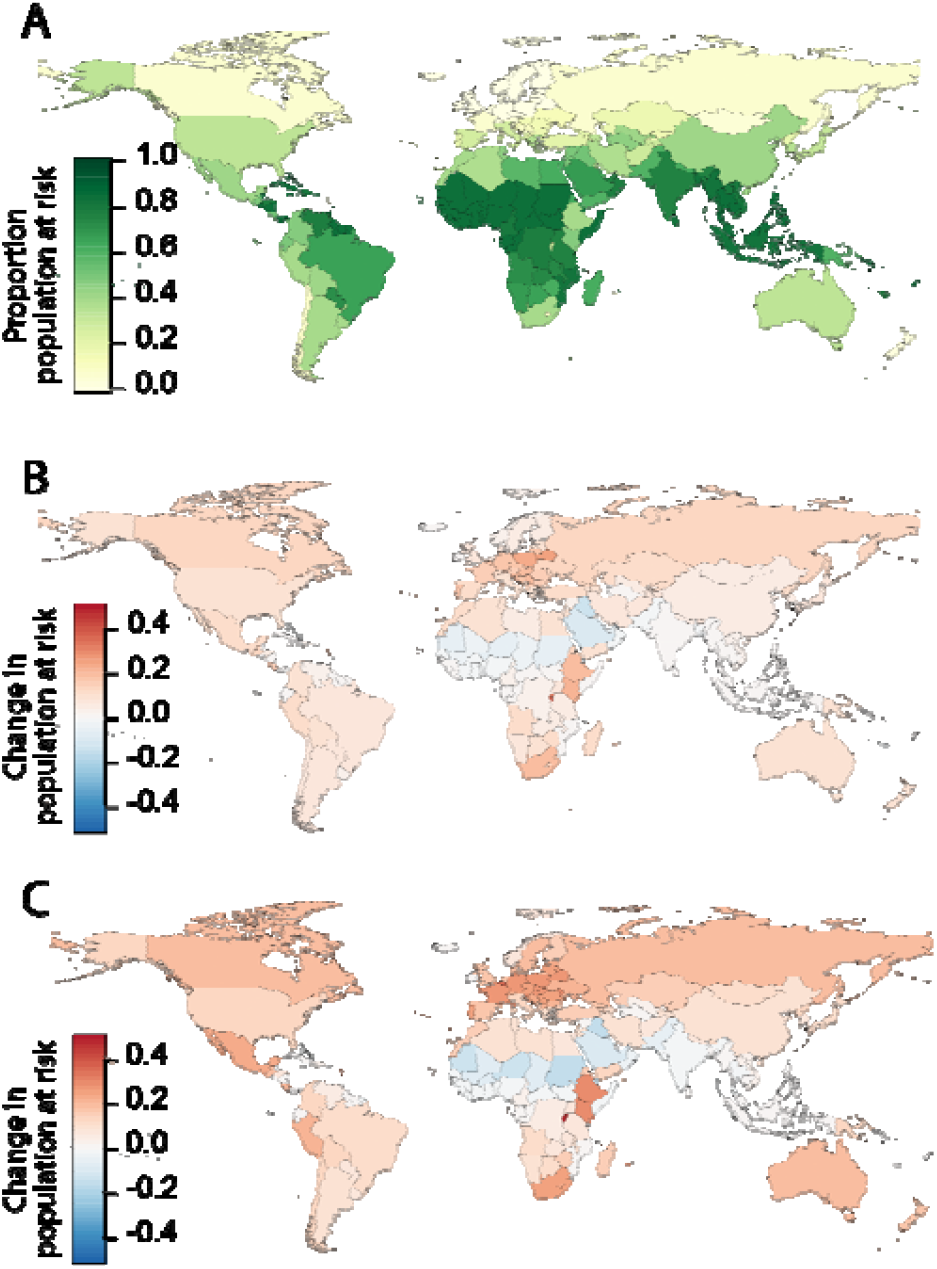
Population at risk. **(A)** Estimated proportion of each country’s population that is at risk of CHIKV infection today. **(B)** Change in the proportion of each country’s population that is at risk of CHIKV infection from today to 2050 under RCP2.6. **(C)** Change in the proportion of each country’s population that is at risk of CHIKV infection from today to 2050 under RCP8.5.

We next estimated the number of infections, cases and deaths in each scenario by country (Figure 3A-B). We assumed that the countries that experience epidemic circulation today (i.e., an average of one outbreak every six years and an average annual force of infection of 1.6% as previously estimated^2^) will continue to experience epidemic circulation, that endemic countries also continue to have endemic circulation (i.e., annual transmission with an average force of infection of 2.1%^2^), and that newly affected countries experience epidemic transmission. Under these assumptions, under RCP2.6, we found that the global annual burden of infections would rise by 35% to 68 million infections per year, cases would increase by 49% to 4.7 million, chronic cases to 2.3 million, with the number of annual deaths rising by 128% to 16,000. Under RCP8.5, the number of infections would increase by 28% to 65 million, the number of cases by 47% to 4.6 million, chronic cases to 2.3 million and the number of deaths by 151% to 17,000. The impact differed substantially by continent, with Europe having the greatest increase (140% increase in cases and 248% increase in deaths under RCP2.6) and Asia the smallest change (29% increase in cases and 117% increase in deaths under RCP2.6) (Figure 3C-D). The share of the global burden was greatest in Asia (54% of all global cases and 63% of deaths under RCP2.6) and Africa (32% of global cases and 14% deaths under RCP2.6) (Figure 3E-F). The impact on case burden was greater under RCP2.6 than RCP8.5 in Africa and Asia, driven by the negative consequences of heat on mosquito survival under RCP8.5, with the burden greatest under RCP8.5 in the other continents. In a sensitivity analysis, we varied the underlying force of infection in epidemic and endemic settings to be either reduced (from e.g., improved living conditions which minimize exposure to mosquitoes) or increased (from e.g., greater global connectivity increasing the frequency of outbreaks). For example, Europe has historically seen smaller outbreaks than elsewhere. We find that a 50% reduction in the force of infection would still result in 88,000 annual cases and 190 deaths per year in Europe, whereas a 20% increase would result in 210,000 cases and 1,100 deaths, as compared to 178,000 cases and 930 deaths under the base case assumptions (Figure S1).

**Figure 3.**
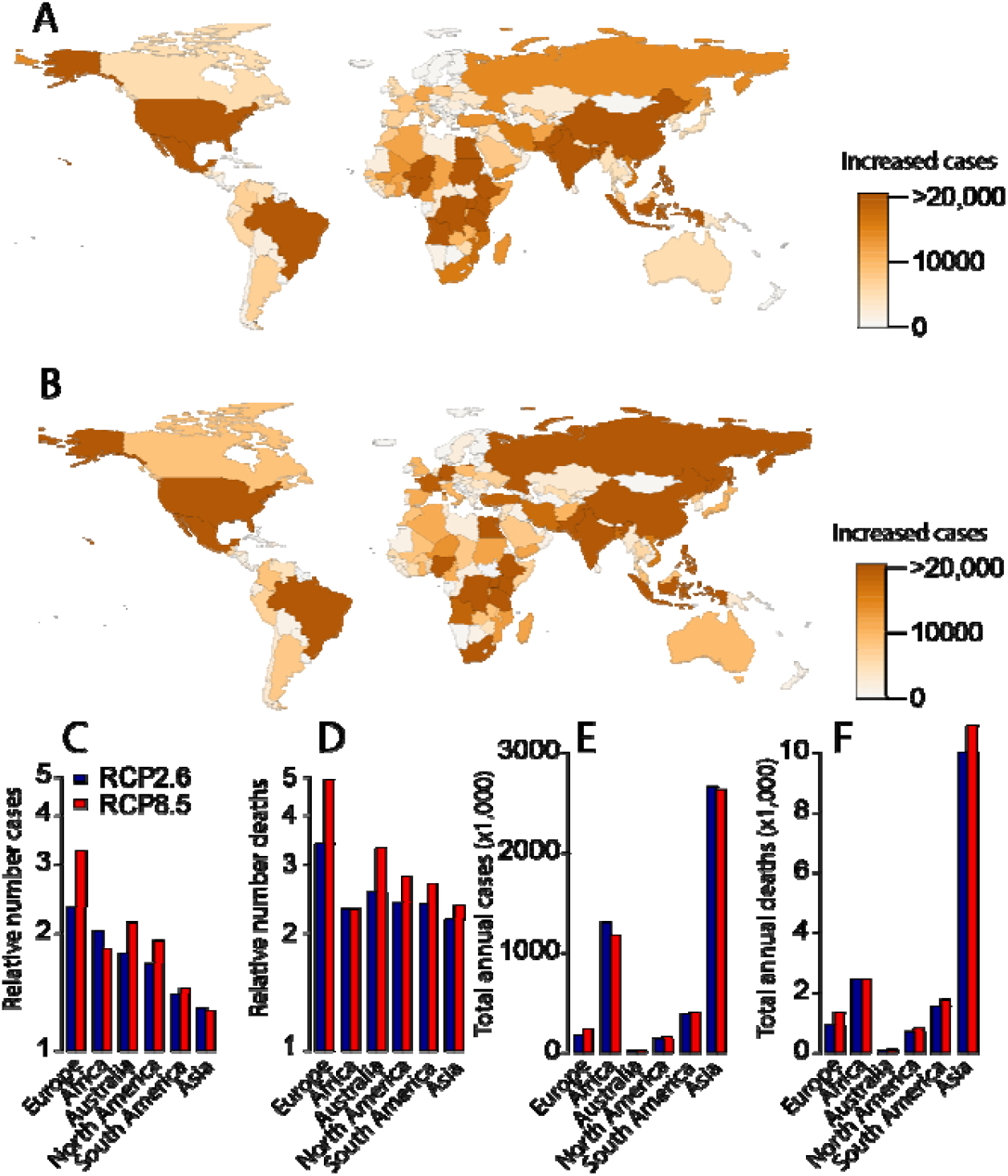
Case burden. **(A)** Difference in absolute number of cases under RCP2.6 in 2025 versus 2025. **(D)** Difference in absolute number of cases under RCP8.5 in 2025 versus 2025. **(E)** Difference in absolute number of deaths under RCP2.6 versus today. **(C)** Relative number of cases in 2050 versus today by continent and RCP scenario. **(D)** Relative number of deaths in 2050 versus today by continent and RCP scenario. **(E)** Absolute number of cases in 2050 by continent and RCP scenario. **(F)** Absolute number of deaths in 2050 by continent and RCP scenario.

### Isolating the independent effects of climate and demographic changes

We found that the independent effect of the expanding presence of the mosquitoes (i.e., without the impact of demography) under RCP2.6 was a 9.0% increase in infections, ranging from 3.7% increase in Asia to 116% increase in Europe, with a similar impact on cases and deaths (Figure 4). The mean independent increase in infections under RCP8.5 was 13.0%. By contrast, the independent effect of population size was a 30.8% increase in infections, a 28.8% increase in cases and a 22.2% increase in deaths. The increase in infections was dominated by Africa (86% increase in infections). Finally, we found that the independent effect of age structure was a 4.3% reduction in infections, a 7.8% increase in cases and a 75.6% increase in deaths.

**Figure 4.**
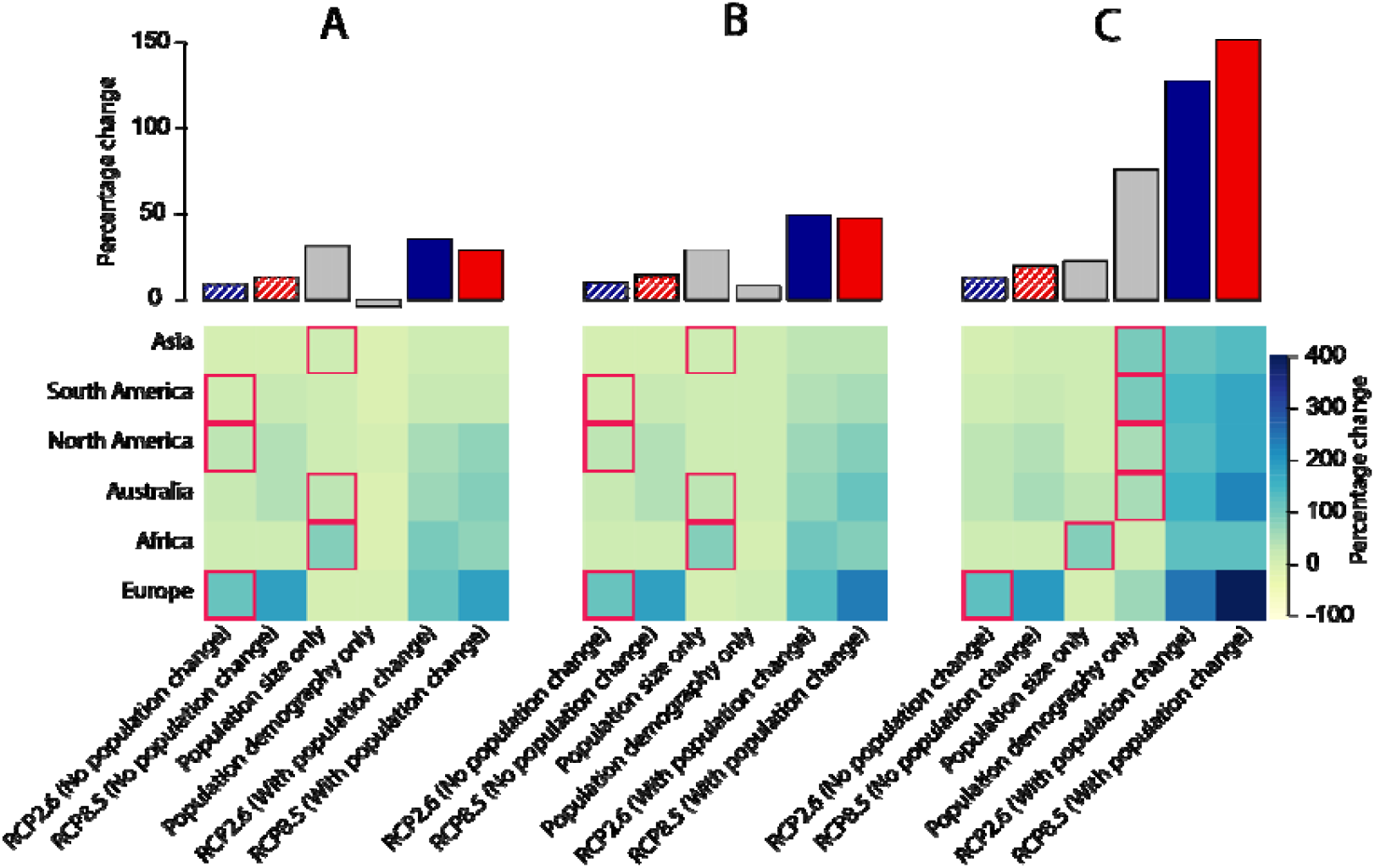
Additional impact of population changes. Impact on **(A)** infections, **(B)** cases, **(C)** deaths under the independent effects of climate changes, the changing population size in 2050 versus today under SSP2 (with the age structure remaining the same as today), the changing age structure of the population in 2050 under SSP2 (with the size of the population remaining the same as today). The final two columns represent the combined effects under the two different climate change scenarios. The top row represents the global average estimates, and the bottom row is divided by continent. The red squares identify the dominant driver of increasing burden by continent under RCP2.6.

Globally, our findings show that population size will be the most important driver of the future global increase in infections and cases whereas changes in age structure will drive increased deaths. However, the cause of the increased burden differed substantially by continent. The increasing footprint of mosquitoes was the most important driver of increased infections and cases in Europe, North America and South America, whereas the change in the size of the population was more important in Australia, Asia and Africa. The main driver of increasing deaths was the expanding mosquito footprint in Europe, however, the growing population was more important in Africa and the age structure (i.e., ageing population) more important in the other four continents.

### Potential impact of vaccination

We considered the potential for vaccination to counteract the increasing burden from CHIKV. We assumed the vaccine had 70% protection against disease and 40% protection against infection with 50% vaccine uptake in those 12 years and older. We also assumed that in epidemic settings, there are vaccine campaigns in response to outbreaks with an average delay of 20% of infections having occurred before the campaign is fully implemented, whereas in endemic settings, vaccines are incorporated into the routine vaccination programs on a continual basis. We found vaccination would avert 18% of infections, 29% of cases, and 31% of deaths per year under the RCP2.6 scenario, with a similar proportion averted under RCP8.5 (Figure 5). Vaccination of 50% of individuals 12 years and older in all areas at risk would require 2.5 billion vaccines, driven by the large populations of India and China. If we assumed a higher vaccine efficacy of 98% against disease, the proportion of cases averted rose to 39% and deaths to 41%. Reducing the time to respond to outbreaks, so that only 1% of cases have occurred by the time the vaccine campaign is completed would result in 33% cases and 35% of deaths averted. Increasing coverage to 90% would result in 48% of cases and 51% of deaths averted. Finally, a full integration of the vaccine into the Expanded Programme on Immunization so that 90% of individuals are protected from 1 year of age in both epidemic and endemic settings would avert 89% of cases and deaths. This final scenario is the most efficient in terms of the number of cases and deaths averted by dose but would require the most vaccines, with 5.3 billion doses.

**Figure 5.**
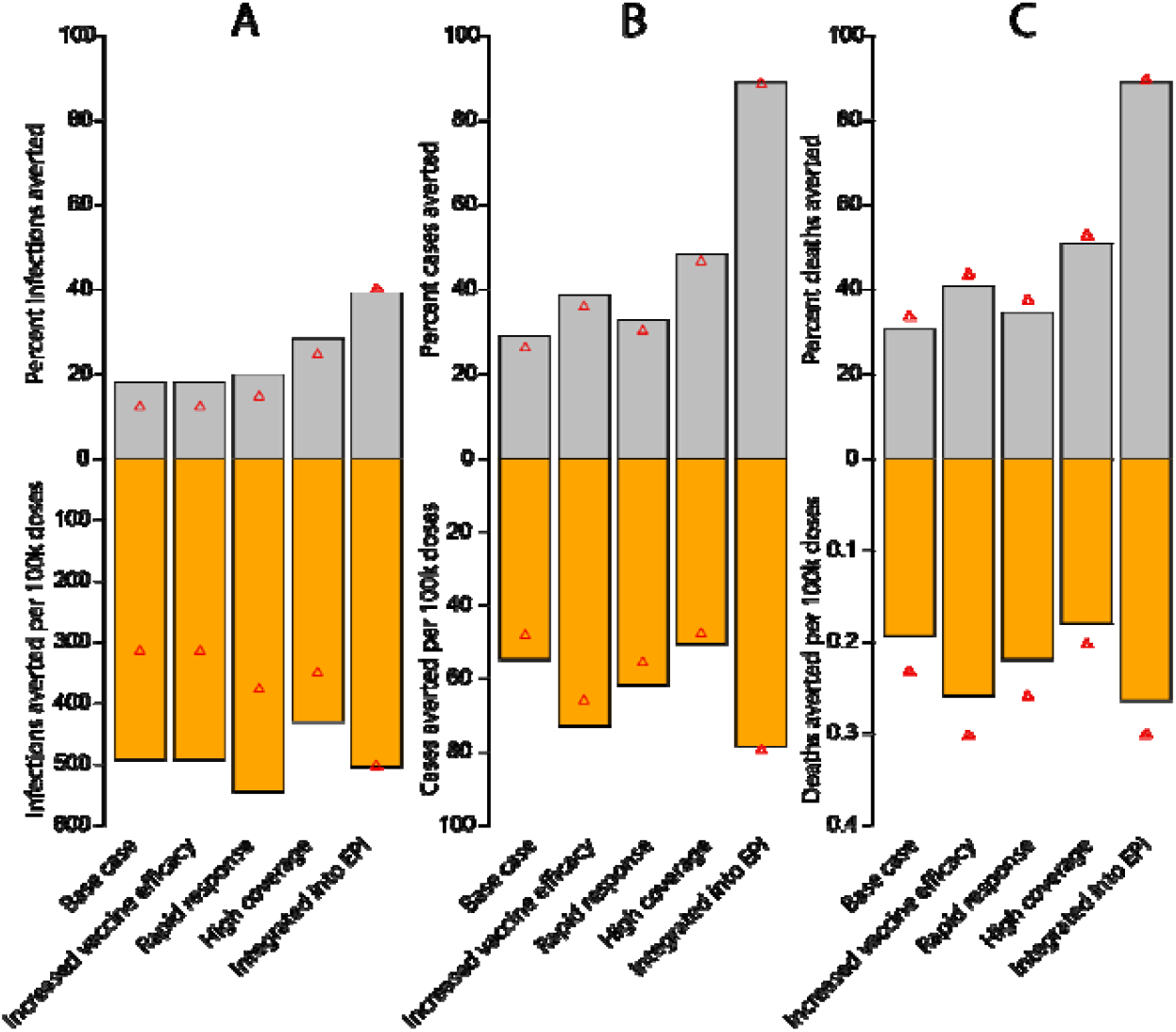
Burden averted through vaccines. **(A)** Infections **(B)** cases and **(C)** deaths averted by year as a percentage (top row) or per 100,000 doses used (bottom row) from vaccination under different scenarios. The bars represent estimates under RCP2.6 and the red triangles from RCP8.5. The base case represents 50% coverage of a 70% effective vaccine against disease (40% against disease) in individuals 12y in age, where in outbreak settings 20% of cases have occurred before deployment is completed. Increased vaccine efficacy represents a 98% effective vaccine. Rapid response means vaccination is complete after 1% of cases have occurred. High coverage represents 90% coverage. Integrated into EPI means 90% of individuals get vaccinated at 1y of age.

## Discussion

We have used the distribution of *Aedes* mosquitoes and population demography projections to quantify the impact of climate change and population changes on CHIKV infection and disease. We found that at a global level there will be a near 50% increase in cases and a more than doubling in the number of annual deaths from CHIKV by 2050. The drivers of this increasing burden differed across the globe, with the expanding mosquito footprint the key driver of infection, case and death burden in Europe and the Americas, whereas the changing demography will drive the burden elsewhere.

Our findings highlight the key role of demography in driving pathogen burden. It has been shown across multiple pathogens, including CHIKV, that older individuals are at increased risk of severe disease and death if infected^20,21^. Protecting vulnerable elderly populations from disease will be critical to minimizing the population burden in at risk regions. This is particularly relevant to countries with older populations, such as in Europe, especially where there has not been substantial historical transmission and therefore there remains high population susceptibility.

We found that climate change will be the principal driver of future CHIKV burden in Europe and the Americas, due to the expansion of the vector and the virus to previously minimally affected countries^22–24^. The more modest impact of climate change in tropical and subtropical regions is largely due to these parts of the world already having mosquito circulation for most of the year and therefore are effectively already at mosquito saturation. Nevertheless, the size of the population in Asia means that this continent will continue to bear the greatest burden from CHIKV, with over half of all cases.

We see modest differences between the optimistic climate change scenario (RCP2.6) and the pessimistic (RCP8.5), which represent two extremes typically used in climate change analysis. These findings suggest that across a wide range of plausible climate scenarios, *Aedes* and CHIKV expansion is largely inevitable. Increasing human travel will lead to more frequent introductions of both mosquitoes and CHIKV virus into previously isolated communities. Currently only a handful of countries across South America, Africa and Asia experience endemic transmission^2^. We could reasonably expect that alongside the broadening footprint of the mosquito, there will be a transition to a greater number of places experiencing more frequent outbreaks. By contrast, improvements in housing and infrastructure are linked to reduced arbovirus burden^25^, suggesting that increased economic development may help mitigate some of the infection risk. Further, in areas dominated by *Aedes aegypti* transmission, the future targeted release of Wolbachia infected mosquitoes may also be an important intervention tool^26^.

We found that vaccines have a critical role in helping counteract CHIKV burden. Vaccinating half of the population aged 12+ years in areas at risk would avert three in ten cases and deaths when we make the assumption that the vaccine only has an efficacy of 70% against disease. As both licensed vaccines were approved using a correlate of protection and not via traditional placebo control phase III trials, we do not have estimates of their vaccine efficacy. However, both vaccines mount high post vaccination titers, which has been strongly associated with protection from infection and disease ^15,27–30^. We could therefore consider that 70% is a conservative estimate. It is important to note that we do not consider the risks associated with vaccination. Adverse events, including a reported death, following Ixchiq vaccination may mean that in some low transmission settings, the risks of vaccination may outweigh the benefits ^31^. The risk from adverse events from live vaccines, such as Ixchiq, are elevated in elderly individuals, who are also the most vulnerable following infection. In the likely scenario that vaccination provides long-lasting protection, introduction of vaccines into populations at younger ages can help these populations as they age, while minimising risk. Such a strategy could also be optimal for emerging settings such as Europe, to help prepare for the growing risk of outbreaks. Introduction of the vaccine into EPI schedules would require further extending the age of eligibility to young children. Age de-escalation studies are currently being conducted in individuals aged 1-11 for both vaccines ^32^

Our estimates are based on modelled estimates of the changing suitability of dengue virus by *Aedes* mosquitoes, and not specifically for CHIKV. The optimal temperature range for CHIKV transmission may be different^33^. However, reassuringly, we find a strong statistical relationship between the *Aedes* estimates and the historical presence of CHIKV outbreaks suggesting that these maps are also informative of CHIKV risk (Figure 1B). A similar strong statistical relationship was obtained using alternative models of *Aedes* suitability^2,7^. Further, our approach implicitly assumes that baseline *Aedes* distribution, and its relationship with CHIKV outbreaks is static, however, *Aedes* populations are continuously changing. The baseline map we used represents an average from 1970-2000, however, in sensitivity analysis an alternative map from 1991-2020 resulted in a consistent relationship with CHIKV outbreaks (Figure S2)^17^, suggesting our baseline estimates of *Aedes* distribution can capture CHIKV risk appropriately. Our estimates also do not account for adaptation of the mosquito to changing environments. It has been suggested that *Aedes aegypti* can adapt to survive increases in temperature and can also adapt behaviourally to seek cooler areas^34,35^. Mosquito adaptation would result in an under-estimate of CHIKV burden, especially in the countries in sub-Saharan Africa and Asia where we estimated a reduction in CHIKV risk from temperature increases. It is also unclear whether viral replicative capacity will be affected by external temperature, although some experimental work has suggested only a moderate impact^36^. Our models assume that the force of infection is identical across affected epidemic (and across endemic) regions. This is a clear oversimplification as the true infection risk will depend on the local environment. However, in the absence of improved understanding of both the current burden and how it will evolve on a more granular level, these crude estimates remain a useful benchmark of future burden. We did not consider intermediate climate change scenarios such as RCP6.0, however, as RCP2.6 and RCP8.5 resulted in largely similar estimates, we can expect that estimates from RCP6.0 would be broadly similar.

Despite these limitations, it is clear that climate change will expand the area at risk for CHIKV outbreaks, especially into new geographies. The health burden will be further heavily increased by demographic changes occurring around the world which will result in older individuals becoming infected, who are more at risk of severe disease and death. Being prepared to counteract this dual threat with vaccines will be central to tackling this growing threat.

## Data Availability

All data used in this article are available online at DOIs 10.1371/journal.pntd.0007213 and 10.1088/1748-9326/11/8/084003

https://journals.plos.org/plosntds/article?id=10.1371/journal.pntd.0007213

https://iopscience.iop.org/article/10.1088/1748-9326/11/8/084003

## Acknowledgements

This project was funded by the Coalition of Epidemic Preparedness Innovations (CEPI). All data used in the paper is publicly available. R Code to recreate the analyses will be made available on GitHub on acceptance.

