## Supplementary Material for "The impact of climate and demographic changes on future chikungunya burden and the potential role of vaccines: a mathematical modelling study"

**Supplementary Figures**

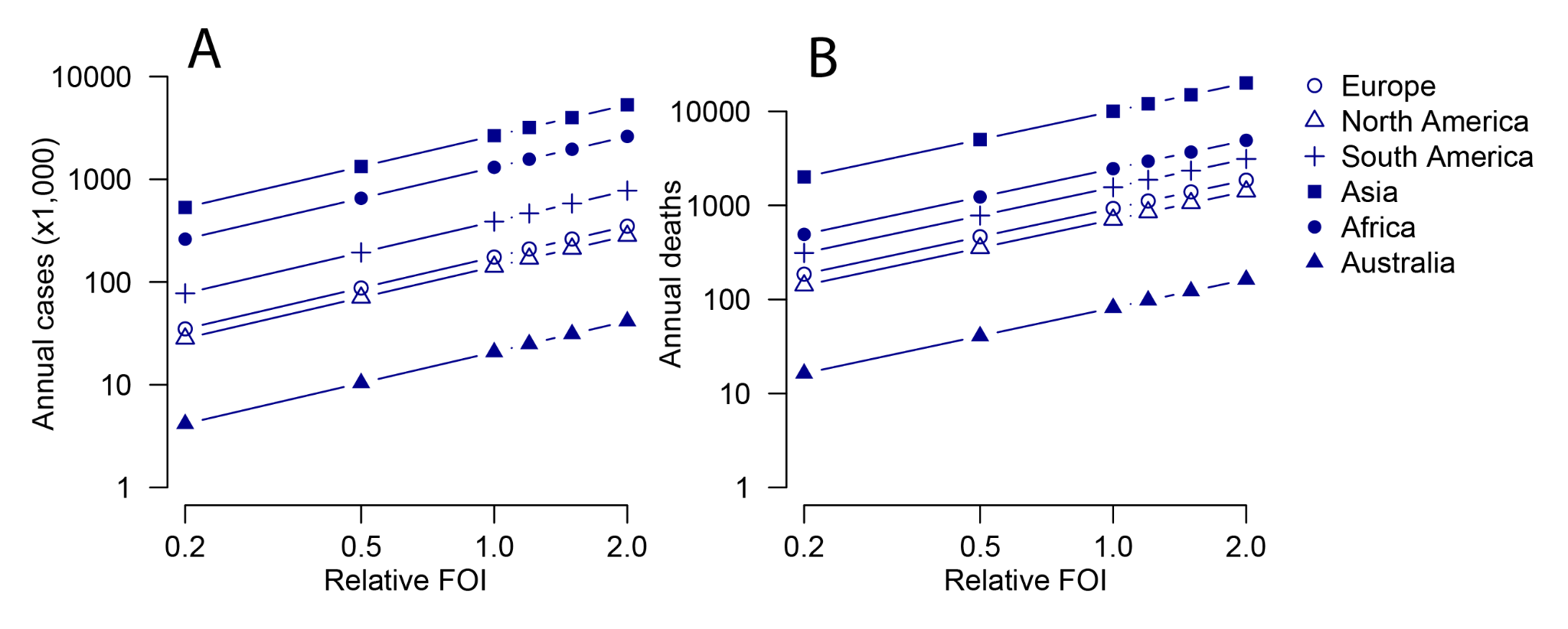

**Figure S1. Sensitivity on the force of infection. (A)** Number of annual cases and **(B)** deaths by continent when the underlying force of infection is altered relative to the base case scenario (2.1% annual force of infection in endemic countries and 1.6% in epidemic countries).

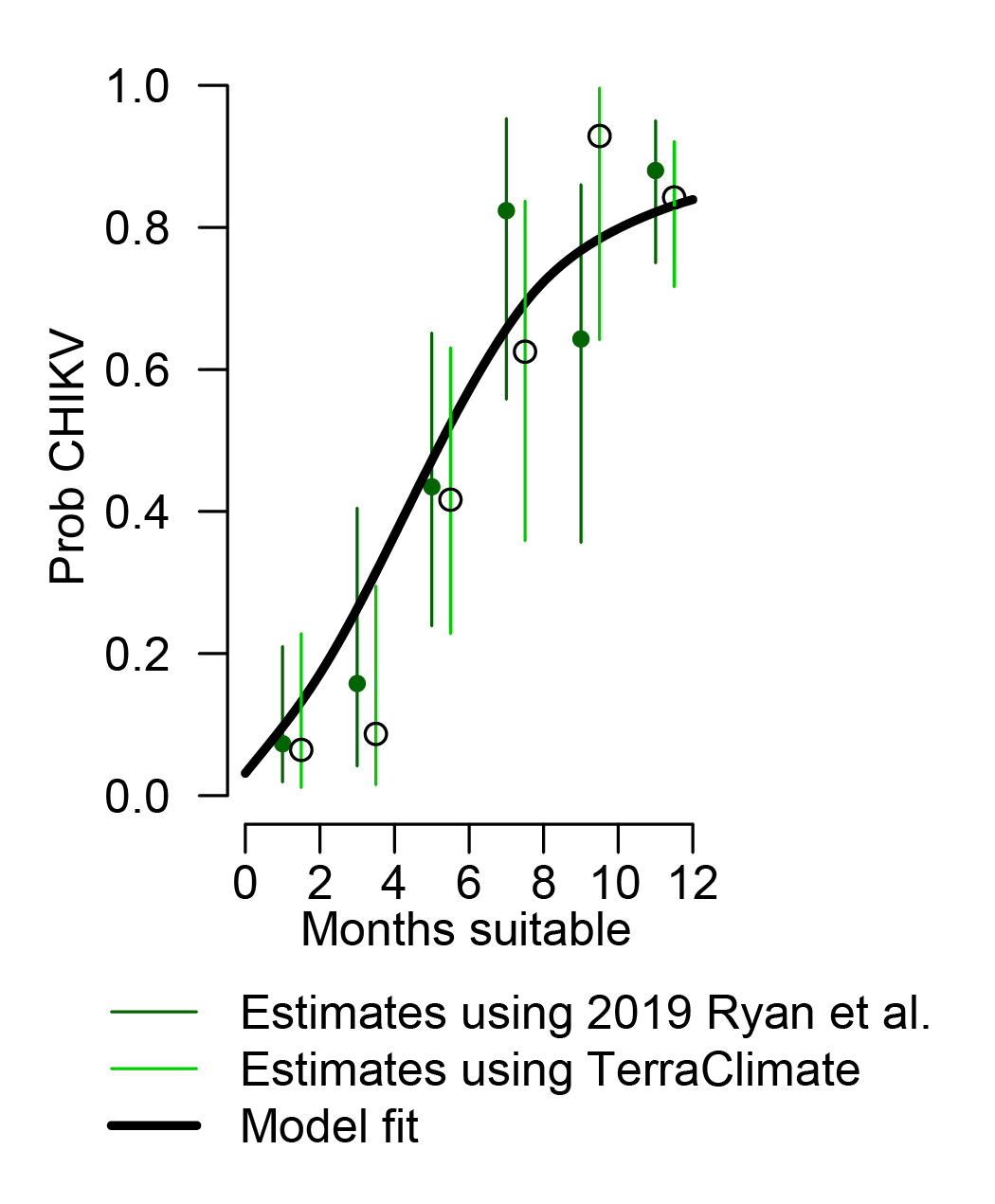

**Figure S2. Sensitivity analysis using a different source of baseline Aedes distribution.** The base case uses the 2019 Ryan et al., estimates^6^. TerraClimate has generated an alternative estimate^17^. Model fit represents the spline model used in the paper.

**Supplementary Text**

**Additional information on the methods**

*Identifying mosquito levels in each country today and under different climate change scenarios*

We use published estimates of the number of months that *Aedes aegypti* and *Aedes albopictus* mosquitoes are present in a year, with each pixel approximately 20x20km^2^ ^6^. Estimates are available for 2020 and for 2050 under the RCP2.6 and RCP8.5 climate change scenarios. To consider the distribution of where human populations will live in 2050, we use modelled estimates of the human population distribution under two different scenarios SSP2 (which we link with RCP2.6) and SSP5 (which we link with RCP8.5) ^16^.

Taking each country with a population size of over 200,000 individuals (UN estimates) in turn, we divided the country into 100 x 100m^2^ grid cells. We then calculated the average total population-time at risk for country j in scenario *k* (today, RCP2.6/SSP2 in 2050 or RCP8.5/SSP5 in 2050) as:

$$MeanMosqBurden_{j,k}=\sum_{i} Pop_{i,j,k}/POP_{j,k}\cdot max(AegyptiMonths_{i,j,k},AlbopictusMonths_{i,j,k})$$

where $Pop_{i,j,k}$ is the human population size in grid cell *i* of country *j* under scenario *k,* $POP_{j,k}$ is the total population in country *j* under scenario *k* and $max(AegyptiMonths_{i,j,k},AlbopictusMonths_{i,j,k})$ is the maximum number of months across the two mosquito species is present in grid cell *i* of country *j* under scenario *k.*

*Linking mosquito levels to CHIKV outbreak risk*

Using the results of a prior literature review, we next explored the relationship between the average mosquito burden within a country (*MeanMosqBurden_j_*) for today and the probability of having ever reported chikungunya transmission in the country. To do this we fit a nonparametric regression model with the smoothing parameter to whether a country has ever experienced chikungunya transmission ^18^:

$$Ever CHIK_{j}=sm(MeanMosqBurden_{j})$$

Where $Ever CHIK_{j}$ represents whether a country has ever experienced CHIKV transmission, as previously identified through a literature review^2^.

*Quantifying the size of the effective population at risk*

Taking each country in turn, we estimated the effective size of the population at risk in each grid cell using the fitted relationship between the mean mosquito burden and national population risk from the repression above. The probability that a resident in country *j* is at risk of infection within a year*,* can be calculated as follows:

$$ProbAtRisk_{j,k}=\sum_{i} Pop_{i,j,k}*tranRisk_{i,j,k}/POP_{j,k}$$

Where $tranRisk_{i,j,k}$ is the estimated probability of transmission in cell *i* within country *j*, calculated using the maximum of *Aedes albopictus* and *Aedes aegypti* months in cell *i* under scenario *k* using the fitted regression model above.

*Quantifying the number of infections, cases and deaths*

The average force of infection in an epidemic setting has previously been estimated at 1.6% per year, while the average force of infection in endemic settings has been estimated at 2.1% per year ^2^. To calculate the probability that an individual of age *a* living in country *j* under scenario *k* gets infected, we used a catalytic model:

$$P(inf{ec}_{a,j,k})=\lambda\cdot exp(-\lambda\cdot min(a,yearsCirculated_{j}))\cdot ProbAtRisk_{j,k}$$

Where $\lambda$ is the force of infection and $yearsCirculated$ is the assumed number of years that CHIKV has circulated in the location.

The number of infections within a country can be calculated as:

$$Ninf{ec}_{j,k}=\sum_{a} P(inf{ec}_{a,j,k})\cdot Pop_{a,j,k}$$

Where $Pop_{a,j,k}$ is the size of the population of age *a* in country *j* under scenario *k* and is obtained from by International Institute for Applied Systems Analysis (IIASA) for SSP2 and SSP5 scenarios ^19^. We run the analysis using single year population sizes but approximate the number of individuals within each age group using estimates of 5-year age bins and dividing by 5. This approach essentially simplifies the model so that each country can be divided into two parts - one part where there is circulation of CHIKV (with either endemic or epidemic circulation) and the remainder with no circulation. While this is a substantial simplification, as we focus on the results at a country level, which will represent an average of these two populations, this is unlikely to be too different from the real average risk of infection in the country, which incorporates all the heterogeneities of infection risk that we could expect across a population.

To capture the number of severe illnesses, chronic infections and deaths following infection, we use the results of a recent study that captured the underlying probability of infection using a seroprevalence study following a large outbreak in Paraguay ^13^.

$$NCases_{j,k}=\sum_{a} P(inf{ec}_{a,j,k})\cdot P(cases_{a})\cdot Pop_{a,j,k}$$

$$NChronic_{j,k}=\sum_{a} P(inf{ec}_{a,j,k})\cdot P(chronic_{a})\cdot Pop_{a,j,k}$$

$$NDeaths_{j,k}=\sum_{a} P(inf{ec}_{a,j,k})\cdot P(death_{a})\cdot Pop_{a,j,k}$$

Where $P(cases_{a})$ and $P(death_{a})$ are taken from the Paraguay study and are available for the following age groups: <1, 1-9, 10-19, 20-29, 30-39, 40-49, 50-59, 60-69, 70+. For *PChronic_a_* we assume that 50% of detected cases have chronic sequelae, consistent with a recent literature review ^37^.

*Isolating the independent effect of demography and climate change*

We estimated the independent effects of the changing distribution of the vector, the changing population size and the changing age structure on infection, case and death burden. To do this, we repeated the analysis but using different subsets of the data. Firstly, we reestimated the burden by country when we used 2025 population estimates but assumed that the spatial distribution of the population at risk was given by RCP2.6 or RCP8.5. We next used the mosquito distribution from today but used the total population size in each country from SSP2 (and separately under SSP5) but keeping the age structure the same as 2025. Finally, we used the mosquito distribution and population size from today but used the age structure from SSP2 (and separately under SSP5).

*Quantifying the impact of vaccines*

To model the impact of vaccines in the transmission scenario where the combined effects of demography and climate change are incorporated, we consider that the force of infection will be reduced due to the partial infection blocking nature of the vaccine.

$$\lambda_{v}=\lambda*(1-\theta)$$

Where $\lambda_{v}$ is the force of infection in a vaccinated population, $\lambda$ is the force of infection in an unvaccinated population and $\theta$ is the effective reduction in the force of infection from the indirect protection generated by the vaccine. From our prior work, we have found that as vaccines are not given to children (who are more likely to be susceptible), under a base case of vaccinating 50% of adults would only result in modest levels of indirect protection, we therefore consider a value of 0.05 for $\theta$.

We then calculated the probability that a vaccinated individual of age *a* in country *j* to be infected as:

$$P(inf{ec^{v}}_{a,j,k})=\lambda_{v}\cdot exp(-\lambda\cdot min(a,yearsCirculated_{j}))\cdot(1-VE_{I})\cdot ProbAtRisk_{j,k}$$

Where $VE_{I}$ is the direct protection from infection provided by the vaccine. This assume that the build up of immunity from natural infection prior to the introduction of the vaccine is dictated by $\lambda$.

The unvaccinated individuals in the same population have the following probability of infection:

$${P(inf{ec^{uv}}_{a,j,k})=\lambda}_{v}\cdot exp(-\lambda\cdot min(a,yearsCirculated_{j}))\cdot ProbAtRisk_{j,k}$$

The overall number of infections in the population then becomes:

$$Ninf{ec^{v}}_{j,k}=\sum_{a} (P(inf{ec^{v}}_{a,j,k})\cdot C_{a}+P(inf{ec^{uv}}_{a,j,k})\cdot(1-C_{a}))\cdot Pop_{a,j,k}$$

Where $C_{a}$ is the vaccination coverage for individuals of age *a*. For epidemic settings, we assume that 20% of infections from that outbreak have already occurred by the time the desired level of vaccination coverage has been achieved.

The number of cases, chronic cases and deaths can then be calculated as:

$$NCas{{es}^{v}}_{j,k}=\sum_{a} P(case_{a})\cdot Pop_{a,j}\cdot(P(inf{ec^{v}}_{a,j,k})\cdot(1-V{E^{*}}_{D})\cdot C_{a}+P(inf{ec^{uv}}_{a,j,k})\cdot(1-C_{a}))$$

$$NChron{{ic}^{v}}_{j,k}=\sum_{a} P(chronic_{a})\cdot Pop_{a,j}\cdot(P(inf{ec^{v}}_{a,j,k})\cdot(1-V{E^{*}}_{D})\cdot C_{a}+P(inf{ec^{uv}}_{a,j,k})\cdot(1-C_{a}))$$

$${{NDeath}^{v}}_{j,k}=\sum_{a} P(death_{a})\cdot Pop_{a,j}\cdot(P(inf{ec^{v}}_{a,j,k})\cdot(1-V{E^{*}}_{D})\cdot C_{a}+P(inf{ec^{uv}}_{a,j,k})\cdot(1-C_{a}))$$

Where $V{E^{*}}_{D}$ is the independent vaccine efficacy against disease, which is calculated as:

$$V{E^{*}}_{D}=(VE_{D}-VE_{I})/(1-VE_{I})$$

This means that the overall protection from the vaccine ($VE_{D}$) represents the total effect of vaccinated individuals that don’t get infected (and therefore cannot get disease) and vaccinated individuals that do get infected but don’t get disease.

We conducted sensitivity analyses where we considered that (a) the vaccine was 98% effective at preventing disease; (b) the vaccine was deployed when 1% of cases were detected (compared to 20% of cases) in responsive campaigns; (c) coverage was 90% and (d) the vaccine was integrated into childhood immunisation so that 90% of individuals over the age of 1years were vaccinated across endemic and epidemic at risk regions.
